# Classifying REM Sleep Behavior Disorder through CNNs with Image-Based Representations of EEGs

**DOI:** 10.1101/2022.04.03.22273365

**Authors:** Saurish Srivastava

**Affiliations:** Evergreen Valley High School, San Jose, CA, USA

**Keywords:** sleep disorders, electroencephalograms, convolutional neural networks

## Abstract

Rapid Eye Movement Sleep Behavior Disorder (RBD) is a parasomnia with a high conversion rate to α–synucleinopathies such as Parkinson’s Disease (PD), dementia with Lewy bodies (DLB), and multiple system atrophy (MSA). The objective of this paper is to classify RBD in patients through a convolutional neural network utilizing imagebased representations of electroencephalogram (EEG) channels.

**Methods:** Analysis was conducted on polysomnography data from 22 patients with RBD and 12 healthy controls acquired from the Cyclic Alternating Pattern (CAP) Sleep Database. EEG channels were split into four frequency bands (theta, alpha, beta, and gamma). Power spectrum density was calculated through a Fast Fourier Transformation (FFT) and converted into 2D images through bicubic interpolation. RBD classification was accomplished through a pre-trained VGG-16 CNN with layer weights being fine-tuned.

**Results:** The model was successful in classifying RBD patients over non-RBD patients and achieved 97.92% accuracy over 40 epochs. Accuracy increased dramatically with increased data generated from FFT and interpolation (62.63% to 97.92%).

**Conclusions:** This project proposes a novel approach toward an automatic classification of RBD and highlights the importance of deep learning models in the field. The proposed transfer learning model outperforms state-of-the-art models and preliminarily accounts for the lack of computational resources in clinical spaces, thereby increasing the accessibility of automatic classification.

**Significance:** By leveraging transfer learning and raw data, the results demonstrate that a similar model for the classification of RBD patients could easily be translated to the clinical atmosphere, drastically accelerating the classification pipeline. The proposed methods are also applicable to α–synucleinopathies, including PD, DLB, and MSA.

## 1. Introduction

Rapid Eye Movement (REM) Sleep Behavior Disorder (RBD) is a prodrome of neurodegeneration that is characterized by a loss of normal muscle atonia and dream-enactment motor activity during REM sleep (Roguski et al. (2020); Högl and Stefani (2016)). RBD is present in about 0.5 to 1.25% of the general population (Haba-Rubio et al. (2017); Kang et al. (2013); Pujol et al. (2017), but given that it is difficult to gauge accurately (Roguski et al., 2020) and because screening is not always conducted, most cases go unrecognized (Atassi et al., 2021). Recent evidence has indicated that RBD is an early predictor for Parkinson’s Disease (PD) (Postuma et al., 2019) – as well as other α–synucleinopathies such as dementia with Lewy bodies (DLB), and multiple system atrophy (MSA) (Stefani and Högl, 2019). At a 14-year follow-up, the “the estimated risk of overt neurodegeneration is around 97% at a 14-year follow-up, while the conversion rate to PD is about Srivastava | | April 3, 2022 | 1–4 90%” (Rechichi et al., 2021). However, rapid development in the drug industry has yielded advancements in new neuroprotective drugs that could only be effective if administered in pre-clinical stages of a pathology (Cruz-Vicente et al. (2021); Devos et al. (2020)). Recognizing RBD early and accurately then becomes invaluable in treatment and prevention of onset and prognosis of future neurodegenerative diseases that may have severe consequences.

Polysomnography (PSG) evidence is the gold standard for diagnosis of RBD, among other sleep disorders and neurological diseases. These PSGs often contain electroencephalography (EEG), electromyography (EMG) and electrooculography (EOG) data, where EEG recordings are frequently used to identify RBD. However, even with ample data, different physicians have been recorded making different assessments about patients (Teplan (2002); Klok et al. (2018); Siuly and Li (2015); Islam et al. (2018)). Thus, there is need to create an automatic classification pipeline that accounts for variation and can learn from past diagnoses. Machine learning is the optimal tool towards approaching this automatic classification problem.

Literature shows numerous studies that utilize conventional machine learning algorithms – such as Support Vector Machine (SVM), *k* -Nearest Neighbors (*k* -NN), and Random Forests (RF) – to classify RBD (Islam et al. (2018); Rechichi et al. (2021); Cooray et al. (2019); Cooray et al. (2021); Buettner et al. (2020)). However, almost all of them utilize massive amounts of features, which can be incredibly computationally expensive. Further, many of these previous approaches do not consider the spatial, temporal, and spectral properties of EEG signals to the fullest extent. Analyzing these channels may be incredibly fruitful to produce a model that achieves high accuracy and maintains robustness. Thus, image-based representations of EEGs, as portrayed in (Arasteh et al., 2021), and their value is important to adequately consider all properties of EEG signals and to maximize robustness (Phan et al., 2019).

The advent of deep learning has deepened interests in applying deep learning to the field of RBD detection. Artificial neural networks, modeled after human neural networks, are powerful in that they can automatically recognize features within data, and deviate from hand-crafted features – which is crucially necessary. In this paper, I propose an automated pipeline for RBD detection through image-based representations, which to the best of my knowledge, has not been explored in this field yet. This novel approach to classify RBD attempts to further improve RBD detection through the power of deep learning, while steering away from conventional machine learning algorithms.

## 2. Materials and Methods

### 2.1. Subjects and Data

Polysomnographic data was obtained from an internationally approved dataset, the Cyclic Alternating Pattern (CAP) Sleep Database (Terzano et al., 2001). Managed by the MIT Laboratory for Computational Physiology on PhysioNet (Goldberger et al., 2000), the CAP database contains a collection of 108 polysomnography recordings of both healthy patients and patients with varying neurological disorders – ranging from bruxism to narcolepsy to RBD. Data from 16 healthy subjects and 22 RBD patients was acquired, however, three healthy subjects were removed due to the presence of ECG artifacts and another two healthy subjects were excluded from analysis due to the absence of EMG recording. Three RBD patients were removed from analysis due to the presence of ECG artifacts. Each recording was subsequently scanned to ensure that at least 5 minutes of REM episodes are present, as handled in (Rechichi et al., 2021).

### 2.2. Pre-processing

The recordings were pre-processed to reduce noise and minimize artifacts. The pre-processing standards used in this paper follow trends established in (Cooray et al., 2019) and (Cooray et al., 2021). All data – EMG, EEG, and ECG – were first resampled at 200Hz. The EEG signal was pre-processed with a 500th order band pass finite impulse response (FIR) filter from 0.3-40Hz. The EMG signal was filtered with a 500th order notch filter from 50-60Hz, as well as a 500th order band pass FIR filter from 10-100Hz. The ECG signal was filtered with a 10th order Butterworth band pass filter between 5-45Hz.

As established in (Lee et al., 2019), data was then bandpass filtered into four conventional EEG frequency bands (Groppe et al., 2013) bands corresponding to wavelength: theta (4–8 Hz), alpha (8–13 Hz), beta (13–30 Hz), and gamma (30–45 Hz). Epoch size was calculated so that there are numerous cycles at the bandwidth’s frequency, resulting in 10-second “mini-epochs.”

### 2.3. Fast Fourier Transformation and Interpolation

Fast Fourier Transformation (FFT) was applied on the time series with windows equal to 1 second to receive 10 different measures per band (10 second mini-epoch sliced into 1 second intervals per band). This resulted in 40 different measures total per subject. These measurements are known as the power spectral density (PSD) estimation of the EEG signal. The Welch’s method was utilized to calculate this PSD, which is given by the following equation: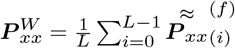.

The measurements for each electrode were then converted into 2D images through a bi-cubic interpolation technique, similar to that of (Fadel et al., 2020). The problem of classification can now be tackled through image recognition. Thus, these images were then fed into a convolutional neural network (CNN).

### 2.4. Model

The power of transfer learning was harnessed to build the model. Transfer learning utilizes a pre-trained CNN on a large dataset, rather than training a CNN from scratch. Although often utilized for when there is not enough data to train an accurate CNN, transfer learning is increasingly being used in other scenarios due to its powerful, comparable results to individually trained CNNs [cite]. Deep CNNs trained on large datasets have the potential to extract features from images that are unrecognizable to the human retina (Ilyas et al. (2019); Geirhos et al. (2020); Simonyan and Zisserman (2014)). Thus, this paper leverages this tremendous power by deploying a pretrained VGG-16 model [cite] on a small dataset to solve the binary classification of RBD patients versus healthy patients. In accordance with the model developed by [cite], this paper modified the base VGG-16 model architecture by utilizing a fully connected layer (1 × 1 × 512) after the last max-pooling layer (7 × 7 × 512), a fully connected layer (1 × 1 × 64) with activation function of “Relu,” and a fully connected layer (1 × 1 × 2) with the activation function of “Softmax” for the classification task consists of two categories, RBD and healthy controls. Figure 2 shows the model architecture for the trained network and Table 2 reports the trained model’s summary.

**Table 1.** Summary of the data utilized in the paper. Comparatively, the healthy subjects are far younger than the patients in the RBD cohort. There are also predominately more males than females in the RBD cohort.

| Cohort | Age | # of Subjects | # of F | # of M |
| --- | --- | --- | --- | --- |
| Healthy | 31.5 ± 4.3 | 12 | 6 | 6 |
| RBD | 70.4 ± 6.4 | 19 | 2 | 17 |

**Table 2.** Trained Model’s Summary

| Layer | Output Shape | Parameter |
| --- | --- | --- |
| vgg16 (Functional) | (None, 7, 7, 512) | 14,714,688 |
| 2D Max Pooling | (None, 1, 1, 512) | 0 |
| Flatten | (None, 512) | 0 |
| Dense | (None, 64) | 32,832 |
| Dense | (None, 2) | 130 |
| Total params: 14,747,650 |  |  |
| Trainable params: 14,747,650 |  |  |
| Non-trainable params: 0 |  |  |

**Figure 1.**
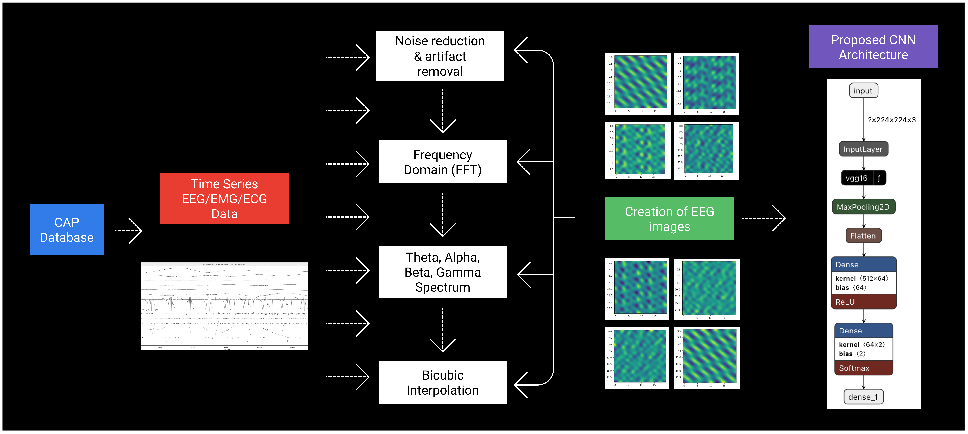
A complete pipeline of the methods

**Figure 2.**
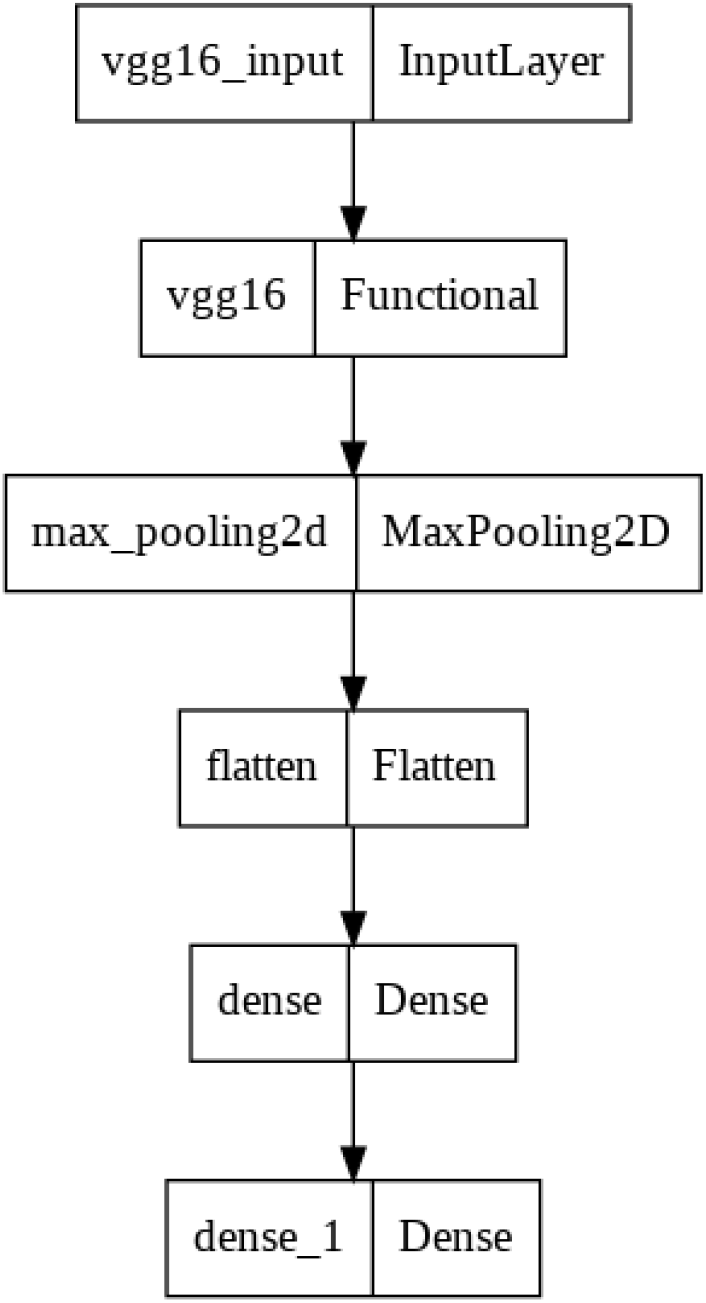
Trained Model Architecture

## 3. Results

CNNs are known to be “data hungry” (Roy et al., 2018), often requiring large amounts of data to achieve high, meaningful accuracy. To explore the effect of a larger dataset, the model was tested four times, each time with a different number of images per patient: 4, 8, 20, and 40. The accuracy for each test are reported in Table 3.

**Table 3.** Comparison of Classification Results with Greater Number of Images

| # of Images per Subject | Accuracy |
| --- | --- |
| 4 | 0.6263 |
| 8 | 0.7576 |
| 20 | 0.9175 |
| 40 | 0.9792 |

As hypothesized, RBD classification increases exponentially with an increase in data. Figure 3 illustrates the training and validation accuracy levels, as well as the corresponding loss per epoch. The final trained model received an accuracy of 97.92% and outperformed all other iterations of the model.

**Figure 3.**
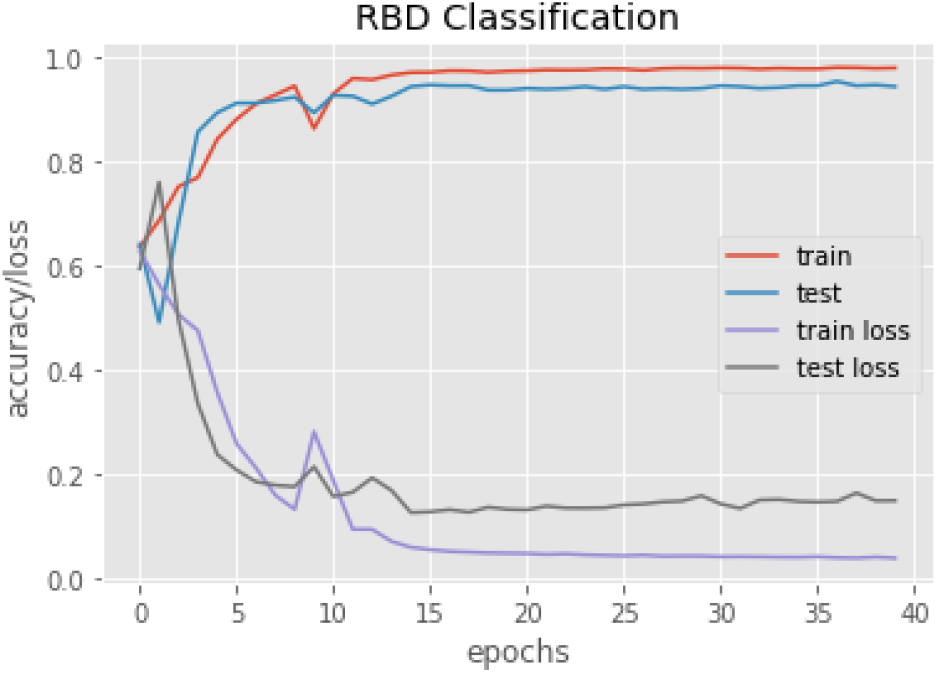
Performance of the model with 40 images per subject

The proposed model outperforms the state-of-the-art models and demonstrates the potential of deep learning in the field of RBD. (Cooray et al., 2019) created a Random Forest (RF) model with an accuracy of 92% through automatic sleep staging and 96% through manual sleep staging. (Rechichi et al., 2021) created a *k* -Nearest Neighbors (*k* -NN) model with an accuracy of 86.96%. However, both models utilize deep methods of calculation for certain features (the RF having the most features) which can be computationally expensive and laborious. It also risks evaluating only certain parts of the Raw data, which could be limiting.

## 4. Discussion and Conclusions

### 4.1. Limitations and Future Developments

With the implementation of a novel method in this field, it is important to acknowledge that this paper does not come without limitations. The cohort of data utilized in this study was heavily male dominant and RBD patients were significantly older than healthy subjects. This could have led to overfitting of the model (not just to male or older patients, but the entire dataset itself too). To maintain consistent dimensionality for images, the number of cycles in each epoch may have differed, which could translate to biases in prediction.

Further, due to a lack of sensor data and information, a complete Azimuthal Equidistant Projection (AEP) (Anderson, 1974) and Clough-Tocher interpolation (Alfeld, 1984) could not be conducted as performed in (Fadel et al., 2020). Thus, this preliminary study may not be able to validate the complete power of convolutional neural networks in this field. Additionally, the ICSD-3 criteria for diagnosis of RBD relies on both REM Sleep Without Atonia (RSWA) — lack of regular muscle atonia during REM sleep — as well as dream enacting and violent behavior during REM sleep, which is reported through a questionnaire often filled out by a sleeping partner. This paper is unable to automatically assess sleep for this dream enacting. However, this paper still provides ample evidence and reason to explore an automatic classification of RBD through neural networks to better accuracy levels and simplify the process for RBD classification for neurologists.

This is the first work, to the best of my knowledge, that goes beyond standard machine learning algorithmss to consider multimodal neural networks for RBD classification. Future developments in this field should thus explore the usage of convolutional neural networks for detection on a larger dataset. As acknowledged above, the skewed dataset could have resulted in overfitting and there is a need to utilize larger and more diverse datasets with RBD patients and healthy subjects. A concatenation of numerous datasets, such as the MASS (Montreal Archive of Sleep Studies) cohort, the CAP Sleep Database, and other private datasets, could create this potential, large dataset. However, this dataset could still face a problem that is relevant to this paper: sensor positions should also be taken into consideration to utilize the AEP and Clough-Tocher interpolation techniques. The lack of adequate annotation of different sensors, as well as an unequal number of electrodes present per subject, made it difficult to employ these algorithms. Future developments should try to incorporate these techniques to ensure a closer relationship with the image and the Raw data.

Further, a new CNN model can be designed – one that traverse through each 10 second mini-epoch during REM sleep and calculates only one measurement through FFT. Although this could be both time and computationally intensive, this would ensure that the full Raw data is being utilized, while creating numerous images per patient. There is potential to create more than 40 images, which could improve accuracy. This would also eliminate the potential for an unequal number of cycles at the bandwidth’s frequency, as all the data is being utilized.

### 4.2. Possible Applications to Clinical Atmosphere

Currently, diagnosis of RBD requires a clinical evaluation and presence of REM sleep accompanied with RSWA. However, these processes are laborious and extremely burdensome, and are often too late to adequately prevent the onset of the disorder (St Louis and Boeve, 2017). There is a necessity to not just simplify this process but improve it to ensure that RBD is detected in patients at an early stage. This paper provides a framework for the automatic classification of RBD through polysomnogram (PSG) recordings alone, which would drastically speed up diagnosis. Its low computational cost would also be beneficial for applications to clinics that may not have the computational power to channel through numerous models with billions of endpoints. Further, given that there is high accuracy, patients, if experiencing violent sleep problems, have the potential to get screened for RBD instantly which would be extremely valuable.

Although PSG recordings are heralded as the gold-standard sleep assessment technique, they are impractical for usage outside a laboratory or clinic, due to “its relatively high cost, the specialized training, and the time burden required to conduct and interpret studies” (Chinoy et al., 2020). Sleep-tracking devices have already been developed for consumer usage and attempt to cut costs, while providing ample data for interpretation. Given the data that these devices output, the framework proposed in this paper can also be utilized and determine whether a patient is exhibiting RBD-like phenotypes during their sleep. With the advent of ‘digital’ meetings for doctors – especially during the COVID-19 pandemic – there is strong potential for these devices to be delivered to patients, and upon adequate testing at home, diagnosis can be sped up without the need for the patient to endure burdensome PSG testing at a clinic.

## Data Availability

All data produced are available online at https://physionet.org/content/capslpdb/1.0.0/.

